# Workflow Intervals andOutcomesof Endovascular Treatment for Acute Large-Vessel Occlusion During On- Versus Off-Hours in China The ANGEL-ACT Registry

**DOI:** 10.1101/2021.06.10.21258678

**Authors:** Yunlong Ding, Feng Gao, Yong Ji, Tingting Zhai, Xu Tong, Baixue Jia, Jian Wu, Jiaqi Wu, Yanrong Zhang, Can Wei, Wenjuan Wang, Jue Zhou, Jiali Niu, Zhongrong Miao, Yan Liu, on behalf of the ANGEL-ACT Study Group

## Abstract

**Background:** Acute ischemic stroke (AIS) leads to a substantial burden of disease among the elderly. There may be a delay in or a poor outcome of endovascular treatment (EVT) among AIS patients with large-vessel occlusion (LVO) during off-hours. By using a prospective, nationwide registry, we compared the workflow intervals and radiological/clinical outcomes between patients with acute LVO treated with EVT presenting during off- and on-hours.

**Methods:** We analyzed prospectively collected Endovascular Treatment Key Technique and Emergency Work Flow Improvement of Acute Ischemic Stroke (ANGEL-ACT) data. Patients presenting during off-hours were defined as those presenting to the emergency department from Monday to Friday between the hours of 17:30 and 08:00, on weekends (from 17:30 on Friday to 08:00 on Monday), and on national holidays. We used logistic regression models with adjustment for potential confounders to determine independent associations between the time of presentation and outcomes.

**Results:** Among 1788 patients, 1079 (60.3%) presented during off-hours. The median onset-to-door time and onset-to-reperfusion time were significantly longer during off-hours than on-hours (165 vs 125 minutes, *P*=0.002 and 410 vs 392 minutes, *P*=0.027). However, there were no significant differences between patients presenting during off- and on-hours in any radiological/clinical outcomes (mRS score: 3 vs 3 points, *P*=0.204; mortality: 15.9% vs 14.3%, *P*=0.172; successful reperfusion: 88.5% vs 87.2%, *P*=0.579; sICH: 7.2% vs 8.4%, *P*=0.492).

**Conclusions:** Off-hours presentation in the nationwide real-world registry was associated with a delay in the visit and reperfusion time of EVT in patients with AIS. However, this delay did not lead to worse radiological/clinical outcomes.

**Registraton:** URL: https://www.clinicaltrials.gov; Unique identifier: NCT03370939.

## Introduction

Stroke is the leading cause of death and disability in China^1^. Ischemic stroke accounts for 65%^2^ of stroke patients in China, of whom 35-40% have large-vessel occlusion (LVO)^1^.LVO results in a large ischemic area and can cause severe brain damage^3, 4^, leading to high mortality and disability rates^5-7^. As a landmark in the treatment of acute ischemic stroke (AIS) with proximal intracranial LVO^8-12^, endovascular treatment (EVT) has been widely used in real-world clinical practice. However, stroke causes 2 million neurons to undergo apoptosis every minute, and the brain ages 3.6 years per hour^13^. Therefore, performing EVT as soon as possible in patients with LVO is the key to improving the prognosis^14, 15^.

Approximately half of patients present during so-called off-hours, i.e., evenings, nights, weekends, and holidays, and EVT needs to be performed jointly by emergency department staff, digital subtraction angiography (DSA) room nurses, technicians, anesthesiologists, and surgeons. Therefore, off-hours presentation may be associated with a delay in the start of EVT. The MR CLEAN trial found that presentation during off-hours was associated with a slight delay of EVT but that this treatment delay did not translate into worse functional outcomes or an increased rate of complications^16^. Data on off-hours delays in workflow intervals have shown delays in different workflow intervals and related performance, but these data are mostly based on single-center studies and are difficult to interpret because of variations in acute stroke care^17-20^. However, there have been no multicenter studies on the association of off-hours presentation and EVT workflow intervals in Chinese patients with LVO.

The Endovascular Treatment Key Technique and Emergency Work Flow Improvement of Acute Ischemic Stroke (ANGEL-ACT) registry was established to evaluate the utilization and subsequent outcomes of AIS patients who received EVT and has confirmed that favorable outcomes of EVT can be achieved in clinical practice in China^21^. In this study, we analyzed prospectively collected ANGEL-ACT data to observe whether the workflow intervals and radiological/clinical outcomes were different between patients with acute LVO treated with EVT who presented during on-versus off-hours.

## Methods

### Study Participants

Data were derived from the ANGEL-ACT registry. ANGEL-ACT was a nationwide, prospective, observational study of 1793 consecutive adult patients with acute LVO treated with EVT at 111 hospitals from 26 provinces in China between November 2017 and March 2019 (https://www.clinicaltrials.gov; unique identifier: NCT03370939). Detailed information about the ANGEL-ACT registry can be found in a previously published article^21^. Ethics approval was granted by the ethics committees of Beijing Tiantan Hospital and all participating centers. Subjects or their representatives provided written informed consent.

For the present study, patients who adhered to the following criteria were included: (1) age ≥18 years; (2) diagnosis of AIS on computed tomography (CT) angiography confirming intracranial LVO; and (3) initiation of any type of EVT, including mechanical thrombectomy, intra-arterial thrombolysis, stenting and angioplasty. Patients were divided into the on-hours group and the off-hours group based on their presentation time.

### Data Collection and Outcome Measures

Off-hours presentation was defined as presentation to the emergency department (participating centers) from Monday to Friday between the hours of 17:30 and 08:00, on weekends (from 17:30 on Friday to 08:00 on Monday), and on national holidays.

All variables, including demographic data, medical history, vital signs, laboratory and neurovascular imaging results, workflow intervals, and clinical outcomes, were prospectively collected.

The workflow intervals included the door-to-puncture time, onset-to-puncture time, onset-to-door time, door-to-imaging time, onset-to-needle time, puncture-to-reperfusion time and onset-to-reperfusion time. The radiological and clinical outcomes included the 90-day modified Rankin Scale (mRS) score asan ordinal variable, functional independence (defined as a 90-day mRS score from 0 to 2), mortality within 90 days, successful reperfusion (defined as the modified Thrombolysis in Cerebral Infarction [mTICI] score of 2b or 3^22^), and symptomatic intracranial hemorrhage (sICH) within 24 hours according to the Heidelberg Bleeding Classification^23^.

### Statistical Analysis

Statistical analyses were performed using SAS 9.4 (SAS Institute, Inc., Cary, NC). All data are described as the median (interquartile range [IQR]) for continuous/ordinal variables and number (percentage) for categorical variables. The Wilcoxon test was used forcontinuous/ordinal variables, and Fisher’s exact test or the chi-square test was used for categorical variables. A *P*-value of < 0.05 was considered statistically significant.Multivariable logistic regression models were used to determine the independent associations between the time of presentation (on-versus off-hours) and radiological/clinical outcomes, with adjustment for age, pre-treatment with intravenous thrombolysis (IVT), baseline NIHSS score, occlusion site, pre-stroke mRS score, and onset-to-door time.

## Results

### Baseline Characteristics

Among the 1793 subjects included in the ANGEL-ACT registry, 5 were excluded because the admission time was missing, leaving 1788 patients eligible for analyses. In total, 1079 patients (60.3%) presented to the emergency department during off-hours, and 709 (39.7%) presented during on-hours. The baseline characteristics were similar in both groups except for the proportion of transferred patients. More patients in the off-hours group than in the on-hours group were transferred from primary stroke centers (36.98% versus 32.02%, P=0.033) (Table 1).

**Table 1.** Baseline Characteristics (median, IQR/ n, %)

| Items | On-hours<br>(n=709) | Off-hours<br>(n=1079) | *P* |
| --- | --- | --- | --- |
| Age,y | 65(55-73) | 66(56-73) | 0.496 |
| Men | 469(66.2) | 705(65.3) | 0.760 |
| Baseline NIHSS (n=1780) | 16(12-22) | 16(12-21) | 0.320 |
| PremRS (n=1787) | 0(0-0) | 0(0-0) | 0.539 |
| Baseline ASPECTS (n=1773) | 9(7-10) | 9(7-10) | 0.414 |
| SBP (mmHg) | 145(130-160) | 145(131-162) | 0.882 |
| Medical history |  |  |  |
| Hypertension | 411(58.0) | 616(57.1) | 0.732 |
| Diabetes | 118(16.6) | 213(19.7) | 0.106 |
| Hyperlipidemia | 66(9.3) | 93(8.6) | 0.612 |
| Coronary heart disease | 111(15.7) | 162(15.0) | 0.737 |
| Atrial fibrillation | 217(30.6) | 344(31.9) | 0.602 |
| Previous stroke | 159(22.4) | 238(22.1) | 0.862 |
| Smoking (recent or current) | 292(41.2) | 424(39.3) | 0.430 |
| IVT performed | 491(69.3) | 774(71.7) | 0.265 |
| Interhospital transfer | 227(32.0) | 399(37.0) | 0.033 |
| Anesthesia |  |  | 0.384 |
| Local anesthesia only | 322(45.4) | 455(42.2) |  |
| General anesthesia | 274(38.7) | 447(41.4) |  |
| Local anesthesia plus sedation | 113(15.9) | 177(16.4) |  |
| Occlusion site |  |  | 0.912 |
| Internal carotid artery | 185(26.1) | 269(24.9) |  |
| M1 | 298(42.0) | 471(43.7) |  |
| Basilar/ vertebral artery | 149(21.0) | 222(20.6) |  |
| Other | 77(10.9) | 117(10.8) |  |
| Stroke classification |  |  | 0.875 |
| Large atherosclerotic stroke | 344(48.5) | 533(49.4) |  |
| Cardiogenic cerebral embolism | 229(32.3) | 346(32.0) |  |
| Other stroke with definite etiology | 79(11.1) | 124(11.5) |  |
| Stroke of unknown etiology | 57(8.0) | 76(7.0) |  |

2 Total number is 1788, unless otherwise indicated.
3 NIHSS indicates National Institutes of Health Stroke Scale; mRS, modified Rankin Scale; ASPECTS, Alberta
4 Stroke Program Early CT Score; SBP, systolic pressure; IVT, intravenous therapy; M1, first segment of the
5 middle cerebral artery.

### Workflow Intervals

The median onset-to-door time during off-hours presentation was 165 (IQR: 70-295) minutes, which was significantly longer than that during on-hours presentation (125 [IQR: 60-260] minutes, *P*=0.002). The median onset-to-reperfusion time was also significantly longer during off-hours (410 [IQR: 310-561] minutes versus 392 [IQR: 285-546] minutes, *P*=0.027). The door-to-puncture time, onset-to-puncture time, door-to-imaging time, onset-to-needle time and puncture-to-reperfusion time were similar between the two groups (Table 2).

**Table 2.** Workflow intervals (median, IQR)

| Items | On-hours<br>(n=709) | Off-hours<br>(n=1079) | *P* |
| --- | --- | --- | --- |
| Door-to-puncture time (n=1787) | 124.5(81.5-190) | 123(83-175) | 0.164 |
| Onset-to-puncture time (n=1774) | 290(200-431) | 305(218-445) | 0.078 |
| Onset-to-door time (n=1754) | 125(60-260) | 165(70-295) | 0.002 |
| Door-to-imaging time (n=1523) | 15(0-30) | 14(0-28) | 0.345 |
| Onset-to-needle time (n=410) | 160(110-220) | 159.5(119.5-213) | 0.843 |
| Puncture-to-reperfusion time (n=1788) | 80 ( 50-128 ) | 88 ( 55-130 ) | 0.078 |
| Onset-to-reperfusion time (n=1774) | 392(285-546) | 410(310-561) | 0.027 |

### Radiological and Clinical Outcomes

No difference was found in the 90-day mRS score between on- and off-hours presentation (median: 3 vs 3 points, adjusted common OR=0.892, 95% CI=0.748–1.064) (Table 3 and Figure 1). We also found no difference between the two groups in the occurrence of functional independence (adjusted OR=0.892, 95% CI=0.724–1.098). The rates of successful reperfusion, mortality within 90 days and sICH were similar in both groups (Table 3).

**Table 3.** Clinical outcomes (median, IQR/ n, %)

| Items | On-hours<br>(n=709) | Off-hours<br>(n=1079) | Unadjusted OR<br>(95% CI) | Unadjusted<br>*P-value* | Adjusted OR<br>(95% CI) | Adjusted<br>*P-value* |
| --- | --- | --- | --- | --- | --- | --- |
| mRS at 90 d<br>(n=1771) | 3(0-5) | 3(0-5) | 0.942(0.794-1.118) | 0.493 | 0.892(0.748-1.064) | 0.204 |
| mRS (0-2) at 90 d<br>(n=1771) | 306(45.7) | 466(44.7) | 0.960(0.790-1.166) | 0.691 | 0.892(0.724-1.098) | 0.280 |
| Reperfusion rate<br>(TICI 2b-3)<br>(n=1788) | 618(87.2) | 955(88.5) | 1.134(0.850-1.514) | 0.414 | 1.087(0.809-1.462) | 0.579 |
| Mortality at 90 d<br>(n=1771) | 101(14.3) | 172(15.9) | 1.142(0.875-1.490) | 0.347 | 1.214(0.919-1.603) | 0.172 |
| sICH within 24h<br>(n=1695) | 56(8.4) | 74(7.2) | 0.842(0.586-1.209) | 0.352 | 0.878(0.606-1.272) | 0.492 |

mRS indicates modified Rankin Scale;TICI, Thrombolysis in Cerebral Infarction; sICH, symptomatic
intracranial hemorrhage.

**Figure 1.**
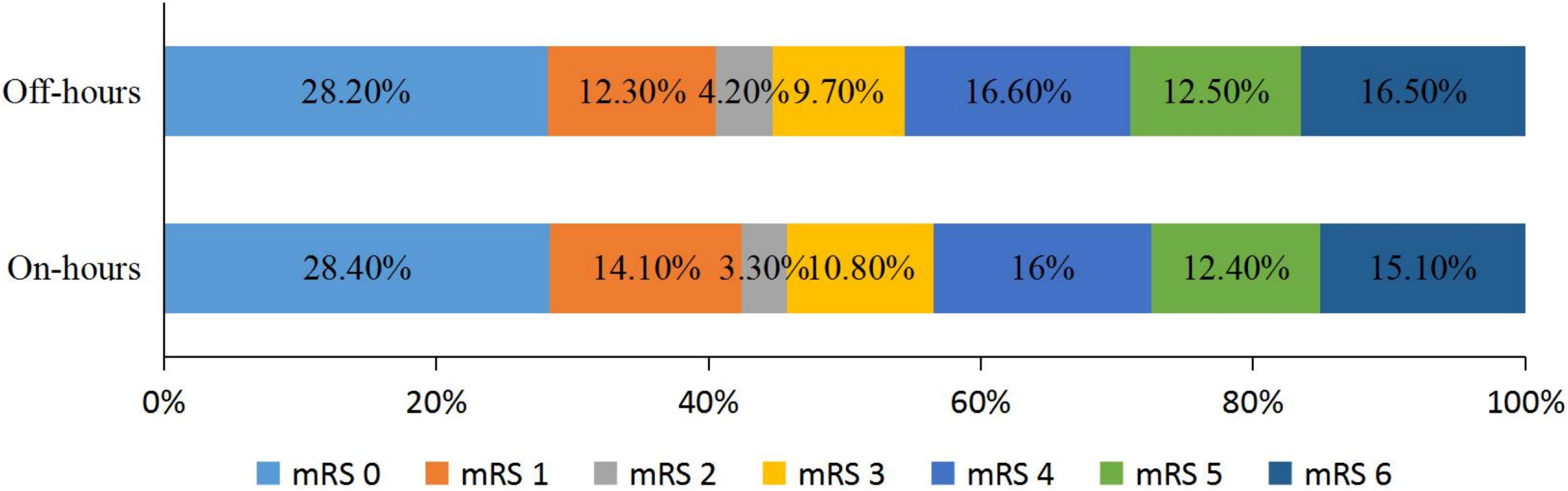
Distribution of modified Rankin Scale (mRS) scores at 90 days.

## Discussion

This large-scale multicenter study reported the relationship of on- and off-hours presentation with workflow intervals and radiological/clinical outcomes and included over 1700 patients treated at 111 different comprehensive stroke centers in China. This study showed that although treatment was administered 40 minutes later in AIS patients presenting off-hours than in those presenting on-hours, there was no difference in the rate of functional independence at 90 days or in the mRS score distribution. In addition, there was no difference in the door-to-puncture time, the rates of successful reperfusion or sICH.

Similar to previous multicenter studies^16, 24-27^, this study did not reveal a difference in the prognosis or rate of complications between the two groups. Benali et al observed a significantly higher rate of good functional outcomes among inpatients admitted at night (51% vs. 35%, P = 0.05)^28^, but another study found a higher mortality rate among patients admitted at night for EVT^29^, which may be due to the heterogeneity of the stroke center process and sample size. Although this multicenter study did not observe an effect of on-versus off-hours presentation on the prognosis, exploring differences in the workflow intervals between the two groups may help to improve the efficiency of EVT implementation. Our research showed that the median onset-to-reperfusion time in off-hours patients was 410 minutes, 18 minutes longer than that in on-hours patients, indicating that there is a certain delay in the EVT process for patients presenting off-hours. The times from onset to door, door to puncture and puncture to reperfusion need to be analyzed to assess the impact of off-hours presentation on the EVT process.

Off-hours patients have a longer onset-to-door time. The 40-minute gap suggests more delays before the hospitalization of off-hours patients. Experiencing a stroke after waking up at night may be an important contributing factor. Another factor may be transfers, as a higher proportion of off-hours patients required transfer. We speculate that some hospitals may not have the capability to provide EVT over 24 hours, so patients who are first diagnosed at these hospitals during off-hours may need to be transferred to a suitable stroke center. Previous research has also described this phenomenon^16^. Notably, in the treatment of patients requiring EVT, interhospital transfer will increase the onset-to-first door time^30-32^; therefore, in establishing the EVT process, the delay caused by referral should be recognized, special attention should be given to the impact of off-hours referrals, and effective publicity should be used to make patients aware of hospitals with 24-hour EVT capabilities in advance so that patients can be delivered directly after stroke onset.

The overall door-to-puncture time in this study was 124 minutes, which is much higher than the 85 minutes reported in a prospective, randomized, controlled study conducted in China^33^ and exceeds the requirement of 90 minutes of advanced Chinese stroke centers, suggesting that the EVT process needs to be further optimized in the real world. However, we observed that the door-to-puncture time was similar in the off-hours group and the on-hours group, which suggests that the EVT process of the stroke center is as efficient off-hours as on-hours and that the emergency green channel, CT examination and DSA are not delayed due to rest. Therefore, this study confirms that presenting off-hours did not cause a delay in the EVT door-to-puncture time in China.

We found that the reperfusion rate in the off-hours group was similar to that in the on-hours group, while the puncture-to-reperfusion time in the off-hours group was 8 minutes longer than that in the on-hours group. We speculate that such a time difference may come from the availability of the off-hours intervention team. According to experience at our center, for patients who require general intravenous anesthesia, the response of the anesthesiologist during off-hours may not be as good as that during on-hours. Furthermore, more junior doctors may be on duty during off-hours, the interventionalists available off-hours may not be as experienced as those available on-hours, and physician or staff fatigue during late-night procedures may cloud judgment or increase the risk of procedural complications^34, 35^. In the future, attention needs to be given to optimizing the configuration of the intervention team during off-hours to reduce the puncture-to-reperfusion time.

The definition of off-hours was set according to statutory holidays and time nodes. The ANGEL-ACT registry contains data from 111 hospitals in 26 provinces in China^21^. These hospitals share Beijing time in Dongba District, which is the standard time in China.Therefore, there is a possibility that when Dongba District has entered the evening, the Eastern Fifth District may still be during the day, so it is necessary to carefully define off-hours. We analyzed the locations of the 111 hospitals and found that 106 (95.4%) hospitals were located in the time zone of the East 8th District and East 7th District. These hospitals accounted for 1716 (95.9%) patients, which means that the time zone difference of 95.9% patients was no more than 1 hour. We conducted a survey on the work and rest time of all hospitals in the group;98.2% of the hospitals’ work hours are 7:30-18:30, with off-hours of 17:00-8:00 in both summer and winter. Therefore, it is reasonable to choose 08:00 and 17:30 when defining the time nodes of off-hours.

This study has some limitations. First, 17 of 1788 patients did not have 3-month follow-up data available from phone interviews; thus, the rates of poor outcomes or serious events may have been underestimated. Second, Saad et al found that the workflow interval had no effect on EVT in teaching hospitals but did have an effect on EVT in non-teaching hospitals^36^. Our study included stroke centers with 24-hour EVT capability, and most of them were teaching hospitals. Therefore, whether the results can be extended to all stroke centers in China still needs further study. Finally, our definition of off-hours included statutory holidays. However, some hospitals included in the study follow a normal work schedule on statutory holidays, and some hospitals even have a 24-hour neurointervention emergency team on duty. This may be the reason why the prognosis of Chinese patients receiving EVT during on-hours and off-hours was similar.

## Conclusion

In conclusion, according to the nationwide real-world registry, off-hours presentation was associated with a delay in the visitand reperfusion time of EVT in patients with AIS, but this delay did not lead to worse radiological and clinical outcomes. In future optimization of the EVT process during off-hours, the onset-to-door time and onset-to-reperfusion time can be key targets for improvement.

## Supporting information

List of ANGEL-ACT study group

## Data Availability

The data that support this study are available from Zhongrong Miao (Department of Interventional Neuroradiology, Beijing Tiantan Hospital, Capital Medical University) upon reasonable request.

## Acknowledgments

We thank all participating hospitals, relevant clinicians, statisticians, and imaging and laboratory technicians.

## Supplemental Materials

ANGEL-ACT Study Group.

