## Supplementary material for "Workflow Intervals andOutcomesof Endovascular Treatment for Acute Large-Vessel Occlusion During On- Versus Off-Hours in China The ANGEL-ACT Registry": List of ANGEL-ACT study group

Beijing Tiantan Hospital, Beijing, China: Zhongrong Miao, MD; Langfang Changzheng Hospital, Hebei, China: Liqiang Gui, MD; Liaocheng Third People's Hospital, Shandong, China: Cunfeng Song, MD; The First People's Hospital of Changzhou, Jiangsu, China: Ya Peng, MD; The Second Affiliated Hospital of Nanjing Medical University, Jiangsu, China: Jin Wu, MD; Fengrun District People's Hospital of Tangshan City, Hebei , China: Shijun Zhao, MD; SiPing Central People's Hospital, Jilin, China: Junfeng Zhao, MD; Yijishan Hospital of Wannan Medical College, Anhui, China: Zhiming Zhou, MD; The 2nd Affiliated Hospital of Harbin Medical University, Heilongjiang, China: Yongli Li, MD; The Central Hospital of Wuhan, Hubei, China: Ping Jing, MD; The First Hospital of Shijiazhuang, Hebei, China: Lei Yang, MD; Shenzhen Hospital of Southern Medical University, Guangdong, China: Yajie Liu, MD; The People's Hospital of Longhua, Guangdong, China: Qingshi Zhao, MD; Jingjiang People's Hospital, Jiangsu, China: Yan Liu, MD; The Third People's Hospital of Hubei Province, Hubei, China: Xiaoxiang Peng, MD; The Second Affiliated Hospital of Guangzhou Medical University, Guangdong, China: Qingchun Gao, MD; Tianjin TEDA Hospital, Tianjin, China: Zaiyu Guo, MD; Zhangzhou Affiliated Hospital of Fujian Medical University, Fujian, China: Wenhuo Chen, MD; Taiyuan Central Hospital, Shanxi, China: Weirong Li, MD; The First Affiliated Hospital of Xinjiang Medical University, Xinjiang, China: Xiaojiang Cheng, MD; Affiliated Drum Tower Hospital of Nanjing University Medical School, Jiangsu, China: Yun Xu, MD; The First People's Hospital of Wenling, Zhejiang, China: Yongqiang Zhang, MD; The Second Affiliated Hospital of Xi'an Jiaotong University, Shaanxi , China: Guilian Zhang, MD; The First People's Hospital of Yulin, Guangxi, China: Yijiu Lu, MD; Zhenjiang First People's Hospital, Jiangsu, China: Xinyu Lu, MD; Qitaihe Coal General Hospital Heilongjiang, China: Dengxiang Wang, MD; People's Hospital of Tangshan City, Hebei , China: Yan Wang, MD; Affiliated Hospital of Guilin Medical University, Guangxi, China: Hao Li, MD; The Affiliated Hospital of Guizhou Medical University, Guizhou Province, China: Yang Hua, MD; The Affiliated Hospital of Xuzhou Medical University, Jiangsu, China: Deqin Geng, MD; Qingdao Central Hospital, ShanDong, China: Haicheng Yuan, MD; The Fourth People's Hospital of Langfang City, Hebei , China: Hongwei Wang, MD; Beijing Daxing hospital, Beijing, China: Haihua Yang, MD; Weifang People's Hospital,ShanDong, China: Zengwu Wang, MD; Luoyang General Hospital Affiliated to Zhengzhou University, Henan, China: Liping Wei, MD; Dongguan Kanghua Hospital, Guangdong, China: Xuancong Liufu, MD; Shunde Hospital of Southern Medical University, Guangdong, China: Xiangqun Shi, MD; Handan Central Hospital, Hebei, China: Juntao Li, MD; The 981 hospital of the Chinese People's Liberation Army, Hebei , China: Wenwu Yang, MD; Linfen people's Hospital, Shanxi, China: Wenji Jing, MD; Anshun people's Hospital of Guizhou , China: Xiang Yong, MD; Changle People's Hospital, Shandong, China: Leyuan Wang, MD; The Second People's Hospital of Dongying, Shandong, China: Chunlei Li, MD; Tangshan Gongren hospital, HeBei , China: Yibin Cao, MD; PLA 985th Hospital of the Joint Logistics Support Force, Shanxi, China: Qingfeng Zhu, MD; Gaomi People's Hospital, ShanDong, China: Peng Zhang, MD; Tongji Hospital, Tongji Medical College, Huazhong University of Science and Technology, Hubei, China: Xiang Luo, MD; Chongqing Sanxia Center Hospital, Chongqing, China: Shengli Chen, MD; Hospital of Traditional Chinese Medicine of Qiannan, Guizhou , China: WenWu Peng, MD; Guangdong Hospital of Chinese Medicine, Guangdong, China: Lixin Wang, MD; People's hospital of Yangjiang， Guangdong, China: Xue Wen, MD; The Third Affiliated Hospital of CQMU, Chongqing, China: Shugui Shi, MD; General Hospital of The Yangtze River Shipping, Hubei, China: Wanming Wang, MD; First People's Hospital of Bijie City,Guizhou, China: Wang Bo, MD; Suqian People's Hospital of Nanjing Drum-Tower Hospital Group, Jiangsu, China: Pu Yuan, MD; Weifang TCM Hospital, Shandong, China: Dong Wang, MD; The Third Affiliated Hospital of Guangzhou Medical University, Guangdong, China: Haitao Guan, MD; Karamay Central hospital, Xinjiang, China: Wenbao Liang, MD; The third people's Hospital of Xinjiang Uygur Autonomous Region, Xinjiang, China: Daliang Ma, MD; Wulanchabu City Central Hospital, Inner Mongolia, China: Long Chen, MD; Hospital of Xinjiang Production & Construction Corps , Xinjiang, China: Yan Xiao, MD; Jiaozuo Second people's hospital, Henan, China: Xiangdong Xie, MD; 904th Hospital of Joint Logistic Support Force of PLA, Jiangsu, China: Zhonghua Shi, MD; Ganzhou People's Hospital, Jiangxi, China: Xiangjun Zeng, MD; 967 Hospital of the Joint Logistics Support Force of PLA, Liaoning, China: Fanfan Su, MD; The Affiliated Hospital of Northwest University Xi'an No.3 Hospital, Shaanxi , China: MingZe Chang, MD; The Second Hospital of Liao Cheng, Shandong, China: Jijun Yin, MD; Jilin Province People's Hospital, Jilin, China: Hongxia Sun, MD; People's Hospital of Huanghua City, Hebei, China: Chong Li, MD; Shanghai Forth People's Hospital, Shanghai, China: Yong Bi, MD; Wanbei Coal-electricity Group Gneral Hospital, Anhui, China: Gang Xie, MD; Shanghai Jiao Tong University Affiliated Sixth People's Hospital, Shanghai, China: Yuwu Zhao, MD; Binzhou Medical University Hospital, Shandong, China: Chao Wang, MD; The 988 hospital of the people's liberation army, Henan , China: Peng Zhang, MD; Linyi People's Hospital, Shandong, China: Xianjun Wang, MD; Yingkou City Central Hospital, Liaoning, China: Dongqun Li, MD; Yantaishan Hospital, Shandong, China: Hui Liang, MD; Mianyang Central hospital, Sichuan, China: Zhonglun Chen, MD; Chengdu Fifth People's Hospital, Sichuan, China: Yan Wang, MD; Hengshui Fifth Hospital of Heng shui City, HeBei, China: Yu Xin, Wang, MD; Second Hospital of Dalian Medical University, Liaoning, China: Lin Yin, MD; Boai Hospital of Zhongshan, Guangdong, China: HongKai Qiu, MD; The First People's Hospital of Yibin, Sichuan , China: Jun Wei, MD; Shanxi provincial people's hospital, Shanxi, China: Yaxuan Sun, MD; Shandong Provincial Third Hospital, Cheeloo College of Medicine, Shandong University, Shandong, China: Xiaoya Feng, MD; Chuxiong State People's Hospital, Chuxiong, Yunnan, China: Weihua Wu, MD; The Fourth Affiliated Hospital of China Medical University, Liaoning, China: Lianbo Gao, MD; Taihe Hospital, Shiyan, Hubei , China: Zhibing Ai, MD; Qingdao Municipal Hospital，Shandong, China: Tan Lan, MD; The First People's Hospital of Yunnan Province, Yunnan, China: Li Ding, MD; The NO.2 People's Hospital of Lanzhou. Gansu, China: Qilong Liang, MD; Taizhou First People's Hospital, Zhejiang, China: Zhimin Wang, MD; Hunan Provincial People's Hospital, Hunan, China: Jianwen Yang, MD; First People's Hospital of Changde City, Hunan, China: Ping Xu, MD; Zhejiang Yuyao People's Hospital, Zhejiang, China: Wei Dong, MD; AideBao Hospital, HeBei , China: Quanle Zheng, MD; The First Hospital of Fangshan District, Beijing, China: Zhenyun Zhu, MD; Tianjin Xiqing Hospital, Tianjin, China: Liyue Zhao, MD; People's Hospital of Zunhua, Hebei, China: Qingbo Meng, MD; Xingtai Third Hospital, Hebei, China: Yuqing Wei, MD; Qingyuan People's Hospital, Guangdong , China: Xianglin Chen, MD; Fengcheng City Central Hospital，Liaoning, China: Wei Wang, MD; People's Hospital of Hejian City, Hebei , China: Dong Sun, MD; Hangzhou Third People's Hospital, Zhejiang, China: Yongxing Yan, MD; Xiangtan Central Hospital, Hunan, China: Guangxiong Yuan, MD; People's Hospital of Nanpi Country, Hebei , China: Yadong Yang, MD; Liuzhou Railway Central Hospital, Guangxi, China: Jianfeng Zhou, MD; Maoming People's Hospital, Guangdong, China: Zhi Yang, MD; Tongde Hospital of Zhejiang Province, Zhejiang, China: Zhenzhong Zhang, MD; The First Affiliated Hospital of Jinzhou Medical University, Liaoning, China: Ning Guan, MD; Xishan coal electricity group worker general hospital, Shaanxi, China: Huihong Wang, MD.
